# Longitudinal Associations Between Endogenous Testosterone, C-Reactive Protein, and Interleukin-6 in Aging Men: Findings from the Baltimore Longitudinal Study of Aging

**DOI:** 10.64898/2026.06.25.26356580

**Authors:** Keerthana Sureshkumar, Meghan R. Grewal, Aaron A. Gurayah, Adam Williams, Justin M. Dubin, Thomas Masterson

## Abstract

**Background:** Elevated C-Reactive Protein (CRP), interleukin-6 (IL-6) and testosterone deficiency are associated with advanced age and chronic inflammatory diseases; while normal testosterone levels have been shown to decrease inflammation through several mechanisms. Cross-sectional studies have shown an inverse relationship between CRP, IL-6 and total testosterone (TT) levels, yet mixed findings have been reported when individual components of metabolic syndrome are considered. We evaluated the relationship between CRP, IL-6 and TT levels in men from 2004-2018 using the Baltimore Longitudinal Study of Aging to determine if low testosterone status is associated with a high inflammatory profile.

**Methods:** Participants were selected from the Baltimore Longitudinal Study of Aging. Male participants with serum TT level measured during at least three visits were included in our cohort. Common measures of inflammatory disease such as CRP, High-Density Lipoprotein (HDL) and Triglyceride levels were collected via blood specimens. Comorbidity data were documented at each visit. Panel regression was used to analyze the relationship of a series of independent variables collected in pooled cross-sectional observations over time with a dependent variable for modeling.

**Results:** A total of 347 patients were included in this study (median age = 70, IQR = 18, average follow up time = 6.7 ± 3.2 years). Participants had a median CRP level of 1.0 mg/dL, median IL-6 level of 3.6, a median TT level of 446 ng/dL. On univariable analysis, increasing TT and HDL levels were associated with a decline in CRP, while high Body Mass Index (BMI), congestive heart failure (CHF), Diabetes, and increased serum triglycerides were associated with increased CRP. Age was not associated with CRP. On multivariable analysis, we found that increasing TT level was associated with a decline in CRP levels, independent of comorbidities (p = 0.018; Table 1). As expected, increased BMI was associated with a significant increase in CRP (p = 0.001, Table 1). Age, CHF, Diabetes, HDL, and Triglycerides were not significant predictors of CRP on multivariable analysis. Similarly, on multivariable analysis, increasing TT levels were independently associated with lower IL-6 levels. Higher HDL cholesterol levels were also associated with lower IL-6 levels, whereas increasing age was associated with higher IL-6 levels. BMI, CHF, diabetes, and triglycerides were not significant predictors of IL-6 (Table 2).

**Conclusions:** Lower levels of serum total testosterone are associated with an increase in CRP in older men over time, independent of chronic inflammatory disease. Given the importance of CRP in pathogenesis of chronic disease, we highlight the potential benefits of using total testosterone as a biomarker of chronic inflammatory states.

**Key findings:**

- In this longitudinal analysis of 347 men from the Baltimore Longitudinal Study of Aging, lower endogenous total testosterone levels were independently associated with higher levels of the inflammatory markers C-reactive protein (CRP) and interleukin-6 (IL-6). These associations persisted after adjustment for factors such as age, body mass index, and major chronic comorbidities. Interestingly, age alone was not independently associated with CRP, while testosterone remained a significant predictor of inflammatory burden over time.

*What is known and what is new?:*

- Prior cross-sectional studies have suggested an inverse relationship between testosterone and inflammatory markers, but findings have been both inconsistent and variable, and it has remained unclear if this relationship reflects aging or the accumulation of chronic disease. Additionally, longitudinal evidence addressing this question has been limited.
- This study provides longitudinal trends indicating that declining total testosterone is independently associated with increases in CRP and IL-6 over time, even after accounting for aging and common comorbidities. The findings suggest that a decline in testosterone is not simply a passive consequence of aging but may be linked to inflammatory processes that extend further than diagnosed chronic disease.

*What is the implication, and what should change now?:*

- These results support total testosterone as a biomarker of systemic inflammation in older men. It suggests that profiling of testosterone levels may offer additional insight into inflammatory risk beyond traditional clinical factors. Our findings support the need for further investigation into the mechanistic relationship between androgen status and inflammation and whether addressing testosterone deficiency could influence inflammation and downstream chronic disease risk in older male populations.

## Introduction

### 1.1 Background

Testosterone is an important hormone that plays a crucial role in the development of male sex organs during organogenesis and drives male puberty during adolescence. Numerous cross-sectional studies have shown an inverse relationship between age and total testosterone, ^1–3, 22^ and longitudinal studies demonstrate a decline in testosterone through the lifespan.^4^ Using longitudinal data collected through the Baltimore Longitudinal Study on Aging (BLSA), Harman et. al report that approximately 20% of men in their 60s and 50% of men in their 80s are hypogonadal.^5^

In the context of increasing life expectancy due to improved medical care, a larger number of men will be advancing further into old age and are at risk of developing clinically significant hypogonadism.

### 1.2 Rationale and Knowledge Gap

However, the underlying pathophysiology of hypogonadism among older men is complicated by the role of chronic inflammation. Though the direction of the relationship is unclear, several studies have established an association between low testosterone and chronic disease such as diabetes, obesity, congestive heart failure (CHF), and even inflammatory bowel disease (IBD) in otherwise young and healthy men.^8–10^ Free testosterone can bind to androgen receptor, a ligand-dependent nuclear transcription factor that is expressed throughout various tissues including muscle, bone, immune, and hematopoietic cells. Therefore it’s been hypothesized that low testosterone can lead to higher levels of proinflammatory cytokines, and vice-versa.^11^

Although several cross-sectional studies have demonstrated inverse associations between testosterone and inflammatory markers, it remains unclear whether these relationships persist longitudinally within individuals as they age and accumulate chronic disease. Longitudinal evidence addressing this question remains limited. Therefore, we utilized repeated measures from the Baltimore Longitudinal Study of Aging to evaluate whether endogenous testosterone levels are independently associated with CRP and IL-6 over time beyond the effects of aging and major comorbid conditions.

As men progress into old age, they are more likely to develop a pro-inflammatory state. and so it’s difficult to determine the extent to which testosterone decline is attributed to the aging process versus the effect of chronic disease.We explored this topic previously by examining the relationship between age, testosterone, and cumulative comorbidities using rich longitudinal data from the Baltimore Longitudinal Study on Aging (BLSA), and found that total testosterone decline is more strongly linked to the presence of comorbidities than age alone.^12^

### 1.3 Objective

Given the lack of evidence elucidating the relationship between aging with chronic illness and natural declines in androgen level, particularly as a longitudinal study, we sought to establish this relationship on a biochemical level by examining the association between testosterone, chronic disease, and pro-inflammatory cytokines CRP and IL-6 among older men using the same 2004-2018 dataset from the BLSA. We hypothesized that a decline testosterone will be associated with an increase in inflammatory markers IL-6 and CRP, independent of age and chronic inflammatory disease state.

### Methods

The men included in this study were selected from the Baltimore Longitudinal Study of Aging (BLSA), a longitudinal cohort study conducted in the United States and supported by the National Institute on Aging of the National Institutes of Health.^20^. Over 3,200 patients have participated in this study since it began in 1958. Participants return to the clinical site every 1-4 years and undergo routine health testing, collection of biometric data, and complete surveys pertaining to general health, medication use, lifestyle etc.

Male participants with documented serum total testosterone measurements were eligible for inclusion in the study. Following application of the study eligibility criteria, the final analytic cohort consisted of 347 participants contributing 698 study visits. Individuals with fewer than three study visits, missing or incomplete data, testosterone replacement therapy use, prostate cancer, or biologically implausible laboratory values were excluded. Following application of these eligibility criteria, the final analytic cohort consisted of 347 participants contributing a total of 698 study visits. Testosterone levels were determined high-performance liquid chromatography-tandem mass spectrometry (Esoterix part of LabCorp, Calabasas Hills, CA, certified by the Centers for Disease Control and Prevention’s Hormone Standardization Project [CDC-HoSt Program]). Measurements of CRP, IL-6, HDL cholesterol, Low-density lipoprotein (LDL) cholesterol, triglycerides, and total testosterone were obtained through the Baltimore Longitudinal Study of Aging (BLSA) protocol. Additional details regarding biomarker collection and laboratory assessment were referenced from established BLSA methodology where available. Measurements of CRP, IL-6, HDL cholesterol, LDL cholesterol, triglycerides, and total testosterone were obtained through the Baltimore Longitudinal Study of Aging (BLSA) protocol. Testosterone levels were determined using high-performance liquid chromatography-tandem mass spectrometry (Esoterix, part of LabCorp, Calabasas Hills, CA, certified by the Centers for Disease Control and Prevention’s Hormone Standardization Project [CDC-HoSt Program]). Additional details regarding biomarker collection and laboratory assessment were referenced from established BLSA methodology where available.

Comorbidity data were obtained from the Baltimore Longitudinal Study of Aging database and included diagnoses of cancer (excluding non-melanoma skin cancer), diabetes mellitus, and congestive heart failure recorded at study visits. These variables were included as clinical covariates in the regression analyses. The BLSA medication database was queried to identify all patients in the cohort who were ever on any form of Testosterone replacement therapy (TRT) during their follow-up period with the BLSA. Individual observations were removed from the analysis for any patients with fewer than 3 total visits, a visit with missing or incomplete data, as well as measured serum testosterone less than 0 ng/dl or greater than 3 standard deviations from the overall mean testosterone level. Similarly, patients with measured IL-6 or CRP less than 0 ng/dl or greater than 3 standard deviations from the overall mean levels were excluded. Patients with documented use of any form of TRT or history of prostate cancer were excluded to eliminate potential confounding alterations in serum testosterone from TRT or androgen deprivation therapy. Missing data were minimal within the final cohort. Of 348 eligible participants, 347 had complete data for all variables included in the analyses. Given the negligible degree of missingness, complete-case analysis was performed and no imputation methods were required.

Statistical analyses were run in R version 4.1.1 (R Core Team, 2021). Descriptive analysis was conducted on characteristics of the final cohort including race, age, follow-up times, total testosterone, CRP, IL-6, BMI distribution, and frequency of comorbidities (Table 1). Categorical variables were represented as frequencies and proportions, while continuous variables were presented as medians with interquartile ranges (IQR). Both univariable and multivariable linear mixed-effects regression analyses were conducted to account for repeated measurements within individuals across follow-up visits. Changes in C-reactive protein (CRP) and interleukin-6 (IL-6) were evaluated as the outcomes of interest. Predictor variables included total testosterone, age, BMI, high-density lipoprotein (HDL), triglycerides, and the clinical covariates congestive heart failure (CHF) and diabetes mellitus (DM), which were included as binary variables. This method of longitudinal panel regression analyses is a statistical technique implemented to analyze the relationship of a series of independent variables collected in pooled cross-sectional observations over time with a dependent variable, and has been used previously in longitudinal data analysis.^13^ This model controls for the effects of unbalanced and/or heterogeneous data (i.e. data collected on subjects at nonuniform timepoints with uneven number of observations).

## Results

A total of 347 patients were included in this study (median age = 70, IQR = 18, average follow up time = 6.7 ± 3.2 years), with a total of 698 visits recorded. Participants had a median CRP level of 1.0 mg/dL, median IL-6 level of 3.60 pg/mL, and median total testosterone level of 446 ng/dL.

On univariable analysis predicting CRP, increasing TT and HDL levels were associated with a decline in CRP, while high BMI and Diabetes, and IL-6 were associated with increased CRP. Age was not associated with CRP. On multivariable analysis, we found that increasing endogenous serum TT level was associated with a decline in CRP levels, independent of age and comorbidities (p = 0.018; Table 2). As expected, increased BMI was associated with a significant increase in CRP (p = 0.001, Table 1). Age, CHF, Diabetes, HDL, and Triglycerides were not significant predictors of CRP on multivariable analysis.

On multivariable analysis, we found that increasing TT, and increased HDL levels were associated with a decrease in IL-6 independent of age and comorbidities (p = 0.03, p = .020; Table 3). Increased age was associated with an increase in IL-6 independent of testosterone and comorbidities (p <.001; Table 3).

## Discussion

### 4.1 Key Findings

Currently, the existing literature does not clarify if the effects aging mediate testosterone levels directly, or indirectly through chronic disease^21^. With the overall increase in life expectancy and increased incidence of inflammatory disease such as metabolic syndrome, hypertension, and diabetes, it’s necessary to understand the biochemical effects of TRT to portend maximum therapeutic benefit.

The principal novel finding of this study is that lower endogenous testosterone levels were associated with higher CRP and IL-6 levels over time in a longitudinal cohort of aging men, independent of age and major chronic comorbidities. Unlike prior cross-sectional investigations, our repeated-measures analysis demonstrates that this relationship persists longitudinally within individuals and is not fully explained by aging alone. Notably, we report a modest association between testosterone decline, increased aging and an increase in IL-6 in this longitudinal sample.

### 4.2 Comparison with Similar Research

Maggio et al. detected a strong inverse relationship between sIL-6r and total T (r = -0.20; P < 0.001) and bioavailable T (r = -0.12; P < 0.05) based on a cross-sectional study of 467 men, aged 65 years or older.^14^ However, they did not report a significant relationship between any other marker of inflammation, including CRP. Though the magnitude of the association was weaker in our study, we were able to replicate this in a longitudinal sample.

We also report an inverse relationship between CRP and testosterone after adjusting for age, and several other potential confounders. Similarly, this relationship has been studied in several cross sectional analyses, and was confirmed by a systematic review by Bianchi et al.^15^ A longitudinal analysis by Osmancevic et al in 2022 demonstrated an inverse relationship between CRP and total testosterone after adjusting for age, waist–hip ratio, hypertension, smoking, type 2 diabetes, and leisure time physical activity (B = −0.26, 95% Confidence interval (CI) −0.41 to−0.11, *P* = 0.001).^16^ However the men included in their study were on average healthier and younger than the men included in our study; additionally, they utilized incongruent methods for measuring total testosterone within their study.

### 4.3 Explanations of Findings

One proposed explanation for the relationship between pro-inflammatory cytokines and TT focuses on the role of free testosterone. Physiologically, 98% of a person’s testosterone is inactive and tightly bound to sex hormone binding globulin (SHBG) or albumin. The remaining 2% of testosterone circulates is free form.^17^ Free testosterone can bind to androgen receptor, a ligand-dependent nuclear transcription factor that is expressed throughout various tissues including muscle, bone, immune, and hematopoietic cells.^11^ A decrease in free testosterone is thought to up-regulate adipokine signaling from fat cells, thus triggering the inflammatory cascade.^18^ The age decline in testosterone can be explained in to varying degrees by decreased production of testosterone by Leydig cells in the testis, an increase in testosterone binding proteins and as well as decreased sensitivity to signaling from the hypogonadal pituitary gonadal (HPG) axis.^19^

### 4.4 Strengths and Limitations

There are several weaknesses in this study worth highlighting. First, this type of multivariate analysis implies an association between two variables but cannot be used to imply causation. While we did detect an inverse relationship between IL-6, CRP and total testosterone we cannot determine the direction of the relationship from this study. Although all variables included in the analyses were selected a priori based on clinical relevance and biologic plausibility, multiple statistical comparisons were performed. Consequently, the possibility of type I error should be considered when interpreting individual statistically significant associations. Second, we included several biomarkers into our models, but were not able to include other important measures such as SHBG, luteinizing hormone, Tumor necrosis factor-alpha (TNF-alpha), etc. Detailed assay-specific information for biomarker measurements was not available for inclusion in the present analysis, as these data were obtained through established BLSA protocols. Additionally, complete longitudinal information regarding lifestyle factors such as smoking status, physical activity, alcohol consumption, and dietary habits was not available for all participants. As these factors may influence both testosterone levels and systemic inflammation, residual confounding cannot be excluded.Third, our study population was mostly white, middle-class, older men, limiting our ability to generalize these biochemical relationships beyond these social factors.

Including these measures would give a better understanding of the mechanism and extent of the observed association. Similarly, we included comorbidity data like the diagnosis of diabetes, CHF and cancer but our study would benefit if we were able to include disease severity, and concurrent treatment status. We could continue to extend our research as this database expands, including men from different racial and social backgrounds. Since there is increasing TRT use in younger patients, it may also be worthwhile to explore if these relationships stand in cohort of younger males.

### 4.5 Implications and Actions Needed

Lower levels of serum total testosterone are associated with a higher level of CRP and IL-6 in older men over time, independent of chronic inflammatory disease. Given the importance of CRP and IL-6 in the pathogenesis of chronic disease, these findings support consideration of total testosterone as a biomarker of systemic inflammation in aging men

## Conclusion

Given the importance of CRP and IL-6 in the pathogenesis of chronic disease, we highlight the potential benefits of using serum testosterone as a biomarker of inflammatory states.

## Data Availability

All data produced are available online at Baltimore Longitudinal Study of Aging.

https://www.blsa.nih.gov

## Acknowledgments

None

## Footnote

### Funding

There are no funding sources associated with this study.

### Ethical Statement

The Baltimore Longitudinal Study of Aging was approved by the National Institute on Aging Institutional Review Board, and all participants provided written informed consent. This analysis used de-identified data.

